# Effects of butyrylated high amylose maize starch (HAMSB) as an adjuvant for oral immunotherapy

**DOI:** 10.1101/2025.09.11.25335560

**Authors:** Duan Ni, Gabriela Pinget, Brigitte Santner-Nanan, Jian Tan, Julen Gabriel Araneta Reyes, Catherine L. Lai, Yanan Wang, Cuong Tran, Julie M. Clarke, Laurence Macia, Dianne E. Campbell, Peter Hsu, Ralph Nanan

## Abstract

**Background:** Oral immunotherapy (OIT) is an important treatment option for food allergy but achieving sustained unresponsiveness (SU) via OIT is challenging. Improving SU for OIT with adjuvants is of great interest but little progress has been made so far. Gut microbiota-derived metabolites like short-chain fatty acids (SCFAs) protect against food allergy in mouse models via promoting regulatory T cells (Treg) generation. We thus aim to investigate the impacts from metabolite-based dietary supplement as an adjuvant in food allergic children receiving peanut OIT.

**Methods:** Based on a prior phase 2 single centre open label interventional randomized controlled trial Oral Peanut Immunotherapy with Short Chain Fatty Acid Adjuvant (OPIA, ACTRN12617000914369), gut microbiota and immune profiles from food allergic children receiving peanut OIT supplemented with butyrylated high-amylose maize starch (HAMSB) or with low amylose maize starch (LAMS) were comprehensively profiled.

**Results:** HAMSB conferred minimal effects on gut microbiota, except transiently increasing their SCFA production like propionate and butyrate. HAMSB skewed CD4^+^FOXP3^+^ Treg towards a tolerogenic phenotype and strikingly increased anti-inflammatory CD4^-^FOXP3^+^ Treg, even after cessation of OIT and HAMSB.

**Conclusions:** We present the first report of the potent immune modulatory effects of dietary butyrate supplementation via HAMSB over an extended period of 1 year in food allergic children. Our findings highlight the tolerance inducing effects of HAMSB and its potential as immunotherapy adjuvant for food allergy and/or autoimmune diseases.

---

Oral immunotherapy (OIT) is an important treatment option for peanut allergy but falls short of reliably achieving long-term tolerance (sustained unresponsiveness, SU). A promising OIT adjuvant candidate for achieving SU is the microbiota-derived short-chain-fatty-acid (SCFA) butyrate. We previously showed that butyrate promotes CD3^+^CD4^+^ regulatory T cells (Treg) development, protecting against severe peanut allergy in a murine model [1]. In this model, butyrate also surprisingly increases CD4^-^FOXP3^+^Treg, (Figure1A-B).

**Figure 1.**
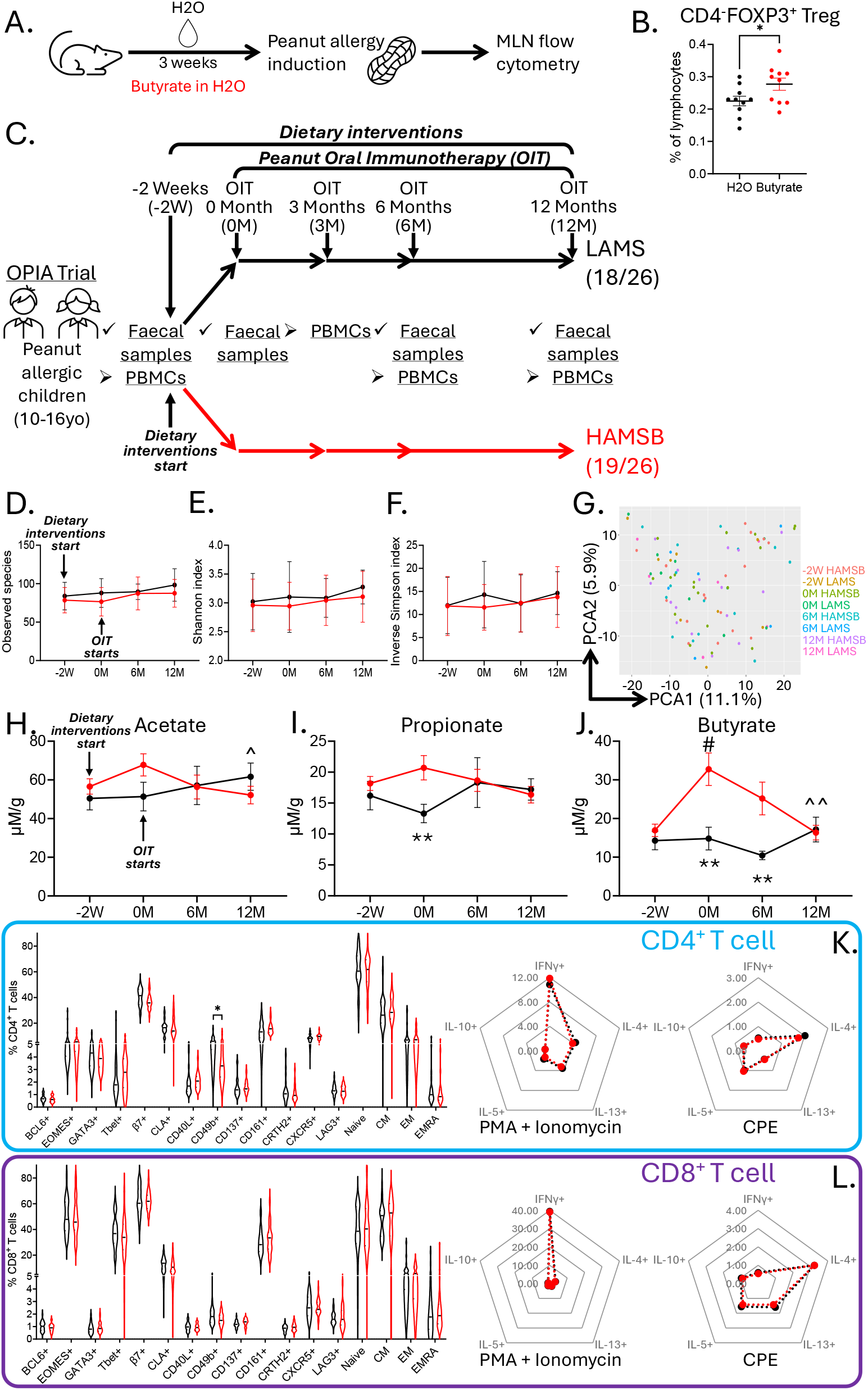
**A**. Overview of peanut allergy mouse model and butyrate treatment. **B**. Butyrate in drinking water treatment increased mesenteric lymph node CD4^-^FOXP3^+^ regulatory T cell (Treg) in peanut allergy mouse model. **C**. Overview of the OPIA trial. **D-F**. Longitudinal changes of observed species (**D**), Shannon index (**E**) and inverse Simpson index (**F**) of LAMS (black) and HAMSB (red) groups in OPIA. **G**. Principal component analysis (PCA) based on Aitchison distance illustrating the microbiota composition of all participants and at all timepoints in OPIA. **H-J**. Longitudinal changes of faecal acetate (**H**), propionate (**I**) and butyrate (**J**) of LAMS (black) and HAMSB (red) groups in OPIA. **K-L**. Immune profiling of CD4^+^ (**K**) and CD8^+^ (CD4^-^, **L**) T cells from LAMS (black) and HAMSB (red) groups in OPIA and their cytokine responses upon stimulations with phorbol 12-myristate 13-acetate (PMA) and ionomycin or crude peanut extract (CPE). Data are represented as mean ± S.E.M. * p<0.05 and ** p<0.01 between groups; # p<0.05 compared with -2W timepoint; and ^^ p<0.01 compared with 0M timepoint.

Encouraged by these observations, the potential OIT adjuvant effects of butyrate supplementation for treatment of human peanut allergy, was tested in the double-blinded, randomized, 3-arm placebo-controlled *Oral Peanut Immunotherapy with butyrate Adjuvant (OPIA*, ACTRN12617000914369) clinical trial [2]. Beyond the published clinical outcomes of the OPIA trial [2], we here focused on 2 arms undergoing OIT with either butyrylated high amylose maize starch (HAMSB) or low amylose maize starch (LAMS) adjuvants (Figure1C). The aim was to dissect the immune and gut microbiome modulatory effects of HAMSB using LAMS as the cross-sectional comparator (Supplementary Information). Clinically, HAMSB and LAMS yielded comparable percentages of participants (18/26 for LAMS and 19/26 for HAMSB) who had successfully achieved the highest dose of daily peanuts in their diets at the OIT 12 Months timepoint (12M), with no difference in SU 6 weeks after intervention cessation [2].

While HAMSB had minimal effects on gut microbiota composition (Figure1D-G), it promoted microbiota derived SCFA production, transiently increasing faecal propionate and butyrate levels at 0M (2-week after dietary interventions, no OIT) and at 6M (Figure1H-J).

Furthermore, HAMSB was associated with reduction of activated CD49b^+^CD4^+^ T cells (Figure1K-L) but had no significant effects on CD3^+^CD4^+^FOXP3^+^Treg frequency (Figure2A, C). In-depth profiling of CD3^+^CD4^+^FOXP3^+^Treg at 12M showed downregulation CD49b and CXCR5 and upregulation of Tbet. Antigen-experienced CD137^+^CD3^+^CD4^+^FOXP3^+^Treg and effector memory CD45RA^-^CD27^-^CD3^+^CD4^+^FOXP3^+^Treg were also increased (Figure2C), indicating skewing towards a tolerogenic phenotype.

**Figure 2.**
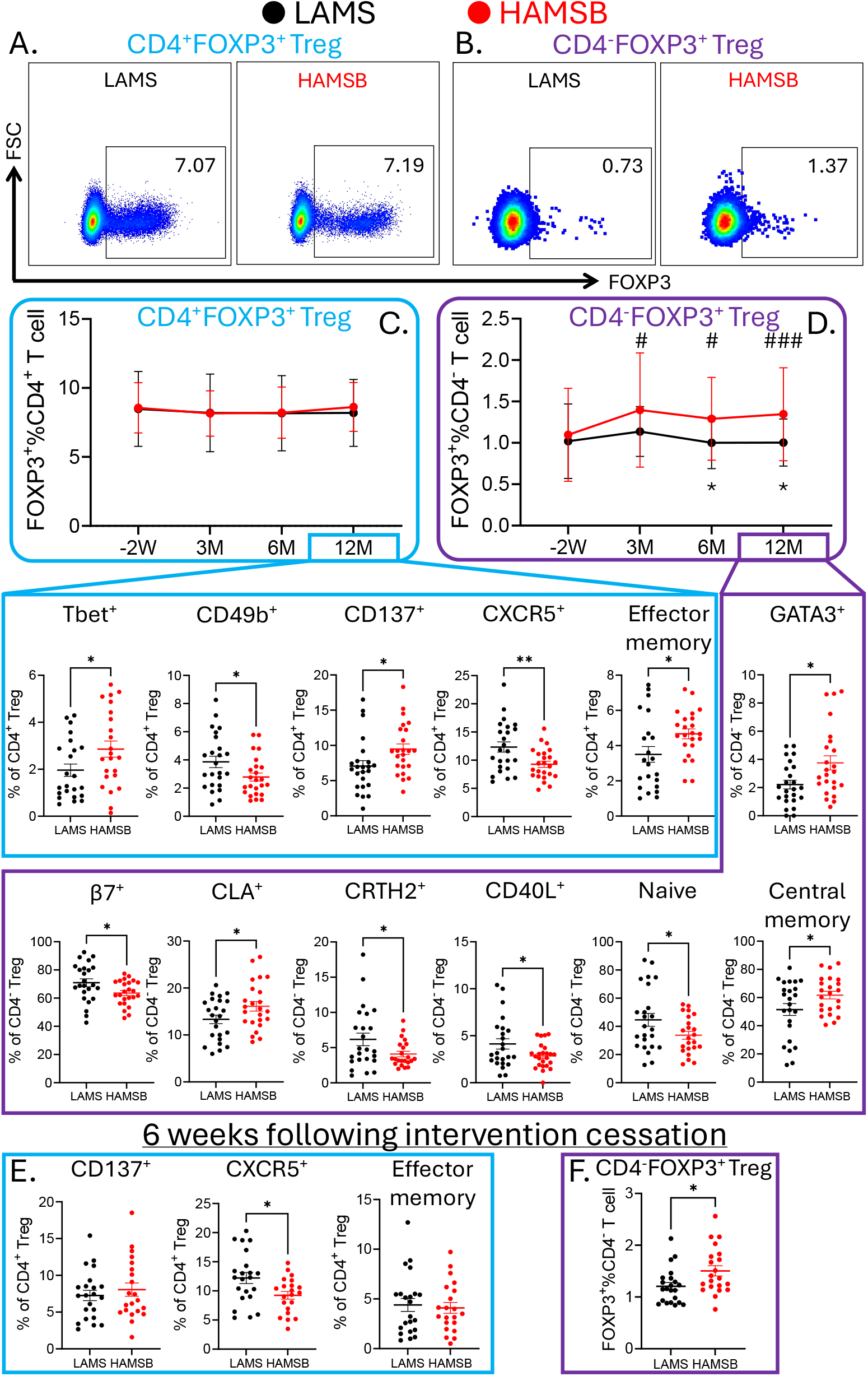
**A-B**. Representative flow cytometric plots of FOXP3 staining in CD4^+^ (**A**) and CD4^-^ (**B**) T cells from LAMS (black) and HAMSB (red) groups in OPIA. **C-D**. Longitudinal changes of CD4^+^FOXP3^+^ (**C**) and CD4^-^FOXP3^+^ (**D**) Treg from LAMS (black) and HAMSB (red) groups in OPIA and changes in their functional subsets at 12M timepoint. **E-F**. 6 weeks following intervention cessation, CXCR5^+^CD4^+^FOXP3^+^ Treg was still higher in LAMS group than HAMSB group but not CD137^+^CD4^+^FOXP3^+^ Treg or effector memory CD4^+^FOXP3^+^ Treg (**E**), while CD8^-^FOXP3^+^ Treg was still higher in HAMSB group than LAMS group (**F**). Data are represented as mean ± S.E.M. * p<0.05 and ** p<0.01 between groups; # p<0.05 and ### p<0.001 compared with -2W timepoint.

Strikingly, similar to our murine findings, butyrate supplementation with HAMSB significantly increased CD3^+^CD4^-^FOXP3^+^Treg (Figure2B) from 3M onwards (Figure2D). At 12M, CD3^+^CD4^-^FOXP3^+^Treg expressed higher skin-homing receptor CLA and lower gut-homing integrin β7. In addition, expression levels of GATA3 were elevated, while CRTH2 and CD40L were reduced. There were also fewer naïve (CD45RA^+^CD27^+^) but more central memory (CD45RA^-^CD27^+^) CD3^+^CD4^-^FOXP3^+^Treg (Figure2D).

6-week after OIT and dietary supplement cessation, CD3^+^CD4^-^FOXP3^+^Treg were still higher in the HAMSB group (Figure1F), indicating a sustained tolerogenic phenotype. In contrast, the moderate effects of HAMSB on CD4^+^ Treg mostly dissipated at this timepoint, except for lower CXCR5 expression levels (Figure1E).

Collectively, this is the first report of the immune and gut microbiome modulatory effects of HAMSB over an extended period of 1 year in humans. The effects on microbiome composition appear to be limited, with a transient increase of microbiota-derived butyrate and propionate production. Strikingly, we reported for the first time that HAMSB increased human CD3^+^CD4^-^ FOXP3^+^Treg and skewed them towards a central memory phenotype with the potential to suppress CD4^+^ effector cells. These effects on CD3^+^CD4^-^FOXP3^+^Treg are significant, as previous reports indicate a similar and important role of CD3^+^CD4^-^CD8^+^Treg as mediators of oral tolerance [3], suppressors of CD4^+^ effector T cell and modulators of antibody production [4]. The fact that HAMSB had different influences on CD3^+^CD4^+^ and CD3^+^CD4^-^ Treg might be explained by butyrate exerting differential effects on these subsets, with respect to epigenetic modifications, processes involving G-protein-coupled-receptor signalling and/or metabolic reprogramming [5].

Despite HAMSB showing no clinical effects as an adjuvant for OIT in the context of the OPIA trial, our analysis provides novel insights into how HAMSB is linked to the expansion of tolerogenic CD3^+^CD4^-^FOXP3^+^Treg. Harnessing the propagating effects of HAMSB on CD3^+^CD4^-^FOXP3^+^Treg could support the development of immunotherapy adjuvant for allergies and/or for autoimmune conditions.

## Data Availability

All data produced in the present study are available upon reasonable request to the authors.

## Author contributions

Study conceptualization and design: R.N., P.H., D.E.C. Investigation and data curation: D.N., G.P., B.S., J.G.A.R., C.L.L., Y.W, C.T., J.M.C., L.M., D.E.C., P.H., R.N. Data analysis: D.N., G.P., B.S., J.G.A.R, C.L.L. Animal experiments: J.T., L.M. Funding and resources: D.C., R.N., P.H. Manuscript preparation: D.N., P.H., R.N. All authors read, edited and approved the final version of the manuscript.

## Acknowledgements

The OPIA trial is funded by a National Health and Medical Research Council Australia Project Grant (NHMRC 1104134). D.N. was supported by the Norman Ernest Bequest Fund.

We acknowledge the contributions of the OPIA study group and help with data management by Katherine Thomson, Matthew Ward, and Ella Ward.

We thank the children and their families for participating in the OPIA trial.

