## Supplementary Information for "Effects of butyrylated high amylose maize starch (HAMSB) as an adjuvant for oral immunotherapy"

**Materials and Methods**

*Mice and allergy model*

Murine food allergy model and butyrate treatment is described in our previous work [1]. In brief, semi-purified AIN93G control diet-fed C57BL/6 mice were given water or 100mM butyrate in drinking water *ad libitum* for 3 weeks. Food allergy was then induced as previously described [1]: Oral sensitisation with 1mg crude peanut extract (CPE) with 10μg of cholera toxin on Day 0 and 7, followed by a booster challenge with 10mg CPE 2 weeks later. Mice were culled 1 week later for analysis by flow cytometry following intraperitoneal challenge with 1mg of CPE.

*Mouse flow cytometry experiment*

Single cell suspension from MLNs was prepared by mechanical disruption with a syringe plunger and filtered through a 100μm cell strainer. Cells were then stained for PerCP anti-mouse CD4 (RM4-5), PE/Cy7 anti-mouse CD25 (PC61) and FITC anti-mouse FOXP3 (FJK-16s). Intracellular staining was performed using the eBioscience™ FOXP3/Transcription Factor kit according to the manufacturer’s instructions.

*Oral Peanut Immunotherapy with butyrate Adjuvant (OPIA) clinical trial*

OPIA was registered on 22 June 2017 in Australian New Zealand Clinical Trials Registry as ACTRN12617000914369. The study was approved by the Human Research Ethics Committee of the Sydney Children’s Hospital Network (HREC/16/SCHN/372). Details about OPIA trial was described in our prior work [2].

*Human faecal short-chain-fatty-acids and microbiota analysis*

Faecal samples were collected as described in our prior work [2]. In brief, all bowel movements over a 48-hour period were collected at 5 timepoints: baseline (-2 weeks, before dietary intervention and OIT initiation), OIT 0 Month (0M, 2-week after dietary intervention and before OIT initiation), OIT 6 Months, and OIT 12 Months.

Faecal SCFA contents were measured using gas chromatography with flame ionization detection (Omega Quant, South Dakota) as previously described [3].

Faecal microbiota analysis was performed commercially by the Australian Genome Research Facility Ltd. similar to our prior studies [4]. Sequencing data was processed and analyzed with R software using *phyloseq*, *microbiome*, and *vegan* packages and results were visualized using *ggplot2* package or GraphPad PRISM.

*Human peripheral blood mononuclear cell sample preparation and flow cytometry experiment*

Peripheral blood mononuclear cell (PBMC) samples were prepared from the OPIA clinical trial and stained as we previously described [2, 5].

*Statistical analysis*

Statistical analysis was performed using GraphPad PRISM. 2-way ANOVA was used for longitudinal comparison and unpaired t-test was run when comparing 2 groups.
